# Comparison of Adverse Effects Related to the Use of Epidural Anesthesia versus Spinal Anesthesia in Cesarean Section Patients at the Plaza de la Salud General Hospital in Santo Domingo, Dominican Republic, During the Period January-December 2021: A Retrospective Study

**DOI:** 10.1101/2023.07.06.23292318

**Authors:** Mariam Alcántara, Alba Rebecca Hernández

## Abstract

**Objectives:** This study aimed to investigate the two different types of regional anesthesia for a cesarean section and to analyze the various perioperative and postoperative side effects related to them.

**Design:** We retrospectively included those patients who required a cesarean section under regional anesthesia

**Setting:** Hospital General Plaza de la Salud from January 2021 to December 2021.

**Participants:** A total of 202 cesarean sections were analyzed. Spinal anesthesia was used in 195 participants and epidural anesthesia in 7; both groups were then compared.

**Results:** Perioperative results revealed that spinal anesthesia had a shorter total anesthesia time, more frequent hypotensive episodes, and a higher perioperative ephedrine administration rate. Regarding the postoperative results, the Apgar scores of the newborns recorded at one minute and at 5 minutes were similar in both groups.

**Conclusions:** For patients who required a cesarean section under regional anesthesia, the use of spinal anesthesia led to a shorter anesthetic time, however it is associated with greater hemodynamic changes and ephedrine administration compared with epidural anesthesia.

## INTRODUCTION

Regional anesthesia is the method of choice for performing cesarean sections, without having the life of the product put at risk and as long as there is no surgical emergency or any contraindication in the patient. In these pregnant patients, regional anesthesia (epidural and spinal) is the most frequently used block technique, since it is associated with greater satisfaction on the part of the mother, fewer hemodynamic changes, fewer systemic changes, and less morbidity. Although the advantages and disadvantages of both anesthesias are widely known, it has not been determined which of these regional techniques has better results. In view of this, it is important to continue addressing these anesthetic methods as an object of study.

Both the epidural and spinal techniques facilitate regional blocks in which local anesthetics are administered in the area adjacent to the spinal cord. These regional anesthesia techniques are preferred over general anesthesia, since these have been shown to have less perioperative and postoperative risk. In addition, they allow rapid extraction of the product of conception in the event of fetal distress.Among the benefits are the rapid recovery of the patient and with less pain than in general anesthesia. In addition, the systemic effects that may occur are preventable.^11^

The center chosen for this research is the Plaza de la Salud General Hospital. In this hospital, anesthesiologists report that the method of regional anesthesia for a cesarean section is chosen following the main international guidelines and based on the skill of the physician who performs the procedure and his academic preparation for each type of block. The different circumstances of each surgery are also taken into account, which include the mother’s consent, contraindications for lumbar puncture, placenta previa, intrauterine depression of the product, among others. Even so, it is an interesting object of study to determine if there are some measurable variables that should be taken into consideration when selecting the anesthetic method to be used in these patients.

Some of the variables to consider include perioperative events such as the rate of anesthetic induction failure, rate of hypotension, the need for intraoperative analgesia, or conversion to general anesthesia after two failed attempts. It is also important to take postoperative events into consideration, such as the neonate’s Apgar score, maternal satisfaction, rate of post-dural puncture headache (which appears from 24 hours and up to 14 days later), postoperative pain, infectious complications such as meningitis or encephalitis, nerve injuries, among others. ^6^ Some investigations have described that spinal anesthesia has shorter anesthetic times and a lower rate of postoperative pain compared to those patients in whom epidural anesthesia is used. However, they do not conclude that there really is one that could be preferred between epidural anesthesia or spinal anesthesia for the use of regional anesthesia. By virtue of this, these investigations suggest that both anesthetic methods should continue to be addressed in different ways as objects of study to assess whether one should be chosen over the other.^6^

The purpose of this research study is to evaluate, through the collection and review of clinical data, the use of epidural anesthesia versus spinal anesthesia in women undergoing cesarean section. Through this, we seek to compare both perioperative and postoperative events that both regional anesthesia techniques may present. As well as specifying which of the two techniques compared in the research is associated with better results in cesarean patients.

### RATIONALE

Obstetric anesthesia refers to anesthetic and analgesic techniques performed during labor, vaginal delivery, cesarean delivery, removal of the placenta, and postpartum tubal ligation.The highest percentage of patients who undergo a cesarean sectionThey do it under regional anesthesia, either spinal or epidural.

Spinal anesthesia is probably the most commonly administered regional technique for cesarean delivery due to its speed of induction and reliability. Epidural anesthesia has a slower motor block and a greater drug requirement to establish adequate sensory block compared with spinal anesthesia. The perceived advantages of epidural block are less possibility of post-puncture headache, slower onset of hemodynamic changes, less possibility of infectious events such as meningitis or encephalitis, and the ability to titrate the local anesthetic through the epidural catheter.^3^

Both techniques are associated with a variety of advantages and disadvantages. However, none of these techniques has been identified as better than the other, nor has it been established whether any are associated with better patient and product outcomes. There are few studies comparing the results of epidural versus spinal anesthesia. At the international level, developed countries such as the United States and England are the ones that have taken the initiative to start a line of research on this topic. Even so, this information is not updated since the most recent study with the greatest scope on this subject is found in the anesthesiology journal “Acta Anesthesiologica Taiwanica” published in 2015.^6^

At the time of carrying out this investigation, the obstetric anesthesia manuals do not prefer the use of one of these anesthetic techniques. Although the advantages and complications of both anesthetic techniques have been described in depth, no effort has been made to identify whether one of these techniques is superior to the other. One of the objectives of this study is to identify the anesthetic technique associated with better patient outcomes. This is important, since the identification of an anesthetic technique associated with a lower rate of complications can lead to better results in patients. This may contribute to the establishment of one of these regional anesthetics as the first line technique in cesarean sections.

In 2019 in the Dominican Republic, 68% of births were via cesarean section, according to the Enhogar-MICS 2019 survey of the National Statistics Office and UNICEF. ^4^ In these pregnant patients, epidural anesthesia and spinal anesthesia are the regional anesthesia techniques most frequently used to perform cesarean sections. In most cases both anesthetic techniques are used interchangeably, often depending on the preference of the anesthesiologist. In the Dominican Republic, like most underdeveloped countries, no studies have been carried out comparing the use of spinal anesthesia versus epidural anesthesia in women undergoing a cesarean section. There is no data that indicates which is used more frequently in the country’s hospitals and clinics, nor in detail which side effects are seen more frequently associated with their use. There have also been no studies that demonstrate which of these anesthetic techniques presents a better prognosis for patients throughout the perioperative and postoperative period.

For the purpose of this research, data were retrospectively collected on the use of spinal anesthesia and epidural anesthesia in women who underwent a cesarean section at the Hospital General Plaza de la Salud during the period January-December 2021. Data on perioperative and postoperative events induced by both anesthetic techniques in these same patients were collected.

### Research Questions

- What is the regional anesthesia technique associated with a lower rate of adverse effects, both in cesarean patients and the product at the Hospital General Plaza de la Salud during the period January-December 2021?
- What are the most common perioperative and postoperative anesthetic events, both in cesarean section patients and the product, with spinal anesthesia?
- What are the most common perioperative and postoperative anesthetic events, both in cesarean section patients and the product, with epidural anesthesia?

### Objectives General Objective

- To compare the side effects related to the use of spinal anesthesia versus epidural anesthesia in women undergoing cesarean section.

### Specific Objectives

- To compare the perioperative events induced by both anesthetic techniques.
- To compare the postoperative events induced by both anesthetic techniques.
- Identify the anesthetic technique associated with better results in patients and the product.

### Limitations

### Theoretical Limitations

- The scarcity of previous studies focused on the use of obstetric anesthesia in the Dominican Republic.
- No studies have been published in the last 5 years comparing the side effects associated with both types of regional anesthesia.

### Practical Limitations

- Incomplete medical records due to lack of continuity in medical care.
- The anesthesiologists’ preference for spinal anesthesia limited us in the amount of data we could collect, since there were few cases in which epidural anesthesia was used.

### Methodological Limitations

- Retrospective data collection limited us to trust that the data provided is accurate.
- Factors that may influence the results such as the degree of urgency, uterine externalization, the experience of the anesthesiologists, the test method for the quality of the block before surgery, as well as the moment and the reason for which anesthetic technique was chosen over the other, were not collected and analyzed in this study.

### Variables

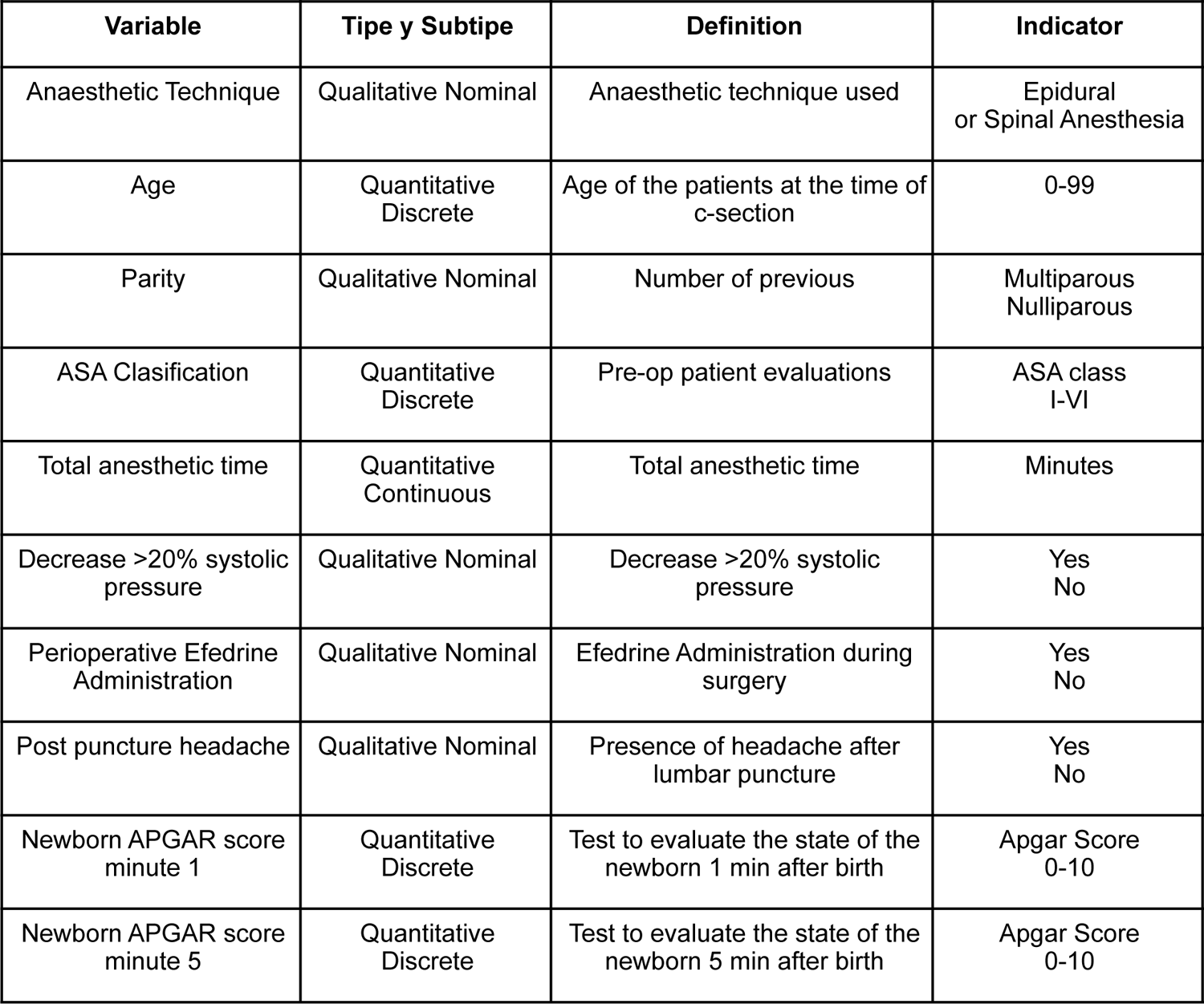

| Variable | Tipe y Subtipo | Definition | Indicator |
| --- | --- | --- | --- |
| Anaesthetic Technique | Qualitative Nominal | Anaesthetic technique used | Epidural or Spinal Anesthesia |
| Age | Quantitative Discrete | Age of the patients at the time of c-section | 0-99 |
| Parity | Qualitative Nominal | Number of previous | Multiparous Nulliparous |
| ASA Clasification | Quantitative Discrete | Pre-op patient evaluations | ASA class I-VI |
| Total anesthetic time | Quantitative Continuous | Total anesthetic time | Minutes |
| Decrease >20% systolic pressure | Qualitative Nominal | Decrease >20% systolic pressure | Yes No |
| Perioperative Efedrine Administration | Qualitative Nominal | Efedrine Administration during surgery | Yes No |
| Post puncture headache | Qualitative Nominal | Presence of headache after lumbar puncture | Yes No |
| Newborn APGAR score minute 1 | Quantitative Discrete | Test to evaluate the state of the newborn 1 min after birth | Apgar Score 0-10 |
| Newborn APGAR score minute 5 | Quantitative Discrete | Test to evaluate the state of the newborn 5 min after birth | Apgar Score 0-10 |

## METHODS

### Context

In pregnant patients, epidural and spinal anesthesia are the most commonly used regional block techniques. Although the advantages and disadvantages of both anesthesias are widely known, it has not been determined which of these regional techniques has better results. (Matos, 2018). The purpose of this research study is to be able to evaluate through the collection and review of clinical data, the use of epidural anesthesia versus spinal anesthesia in women undergoing cesarean section. Likewise, it is hoped to determine which of the two techniques is associated with better results in these patients.

### Modalities of Work

The modality of this study is a research project, based on the scientific methodology for its realization and those results obtained can contribute to professional enrichment both theoretically and practically. This research serves as a local, regional or national contribution in the health sector, specifically in the area of obstetric anesthesia.

### Type of study

This is an observational and analytical study of the retrospective cohort type. Existing data were used to analyze both the exposure to the different types of regional anesthesia in women who underwent a cesarean section, and their results.

### Research methods and techniques

Indirect observation was used as a research method for data collection through clinical records and they were divided into two groups, patients receiving epidural anesthesia and those receiving spinal anesthesia.

### Data collection instrument

Data collection was through the review of clinical records provided by the Plaza de la Salud General Hospital of those patients who underwent a cesarean section during the period January-December 2021.

A simple questionnaire of closed questions was used, answering yes or no and multiple choice questions. The collected data was then deposited in data observation sheets and information collection tables, including the variables that are relevant to the study.

### Ethical considerations

The personal data of the patients who participated in the study, such as the name, address and telephone number will not be published in order to ensure the protection of the rights of human participants who participate in the research and protect their privacy and integrity.

In addition, the research was submitted and approved by the Research Ethics Committee of the Universidad Iberoamericana. It is led by the Dean of Research and Innovation (DEII) and is in charge of evaluating research projects with human beings to ensure that they are carried out responsibly and safely, adhering to international ethical standards.

### Population and sample selection

The population is made up of a representative sample of pregnant women who had an indication for cesarean section at the Hospital General Plaza de la Salud during the period January-December 2021. By calculating the sample size, a group of 202 patients from the population was selected. total of 776 patients. (95% CI, margin of error 5).

The sampling was non-probabilistic at convenience, and a significant sample of women who underwent a cesarean section was obtained at the Hospital General Plaza de la Salud during the period January-December 2021.

### Inclusion and exclusion criteria

#### Inclusion criteria

- Pregnant women over 18 years of age.
- Pregnant women with an indication for caesarean section at the Plaza de la Salud General Hospital during the period January-December 2021.

#### Exclusion criteria

- Pregnant women under 18 years of age.
- Patients with failure in the placement of regional anesthesia.
- Pregnant women with an indication for cesarean section in an institution other than the HGPS.
- Pregnant women with an indication for cesarean section outside the period January-December 2021.

### Procedures for data processing and analysis

Parametric data were presented as mean ! SD (standard deviation). Statistical analysis was performed with Prism GraphPad (Version 9.4.1).

When processing the data, an imbalance was evidenced in them, due to the fact that the majority of the cases were spinal anesthesia. This imbalance is due to the fact that at Hospital General Plaza de la Salud, anesthesiologists prefer the use of this anesthetic method. Data processing and analysis using the most common statistical tests can be affected by an unbalanced data set. The equilibrium problem corresponds to the difference in the number of samples in the different classes. When an unbalanced relationship exists, the results favor the class with the largest number of samples, usually called the majority class.^8^ One approach to address the class imbalance problem is to randomly resample the data set. The primary approach to randomly resampling an unbalanced data set is to duplicate examples from the minority class, which is called oversampling.. The augmented data set should be used instead of the original data set to perform the statistical tests.^1^ However, it is important to note that looking for a balanced distribution for a highly lopsided data set may cause the algorithm to overfit the minority class, which in turn results in increased generalization error.^13^

To compare both groups appropriately, a random oversampling of the minority group. Using this new data, a Student’s t-test and Chi-square test were used to compare continuous and categorical variables, respectively. A p value <0.05 was defined as a significant difference.

## RESULTS

This retrospective study collected data from 202 patients who underwent a cesarean section using one of the two regional anesthesia techniques (epidural or spinal) at the Plaza de la Salud General Hospital during the months of January to December 2021. The final analysis includes 195 patients who received spinal anesthesia and 7 who received epidural anesthesia. The demographic characteristics of age, nulliparity, multiparity and ASA score of both anesthetic techniques were collected. The mean age of the patients was 29.8 ! 5.57 in those who received spinal anesthesia and 26 ! 2.16 in those who received epidural anesthesia. Out of these patients, 26.7% were nulliparous (n=54) and 73.3% were multiparous (n=148). Finally, regarding the ASA classification, 30.2% (n=61) were ASA I, 68.3% (n=138) were ASA II, and 1.5% (n=3) were ASA III.

In the case of perioperative events, it was found that the total anesthetic time in the spinal anesthesia group was 77.1 ! 13.06, and that of epidural anesthesia was 85 ! 10.41 minutes. Hypotensive episodes in the spinal anesthesia group were seen in 63.0% of patients (n=123) versus 42.8% (n=3) of patients in the epidural anesthesia group. Similarly, the administration of ephedrine was seen in 61.5% (n=120) of patients with spinal anesthesia, and in 42.8% (n=3) of patients with epidurals.

Regarding the postoperative effects, 6 cases of post-puncture headache belonging to the group that received spinal anesthesia were identified, showing an incidence of 3.07%. While in the epidural anesthesia group none was noted. In addition to this, the APGAR scale values were collected in the neonates for the spinal and epidural group in the first minute (7.81 ! 0.63 vs. 7.85 ! 0.37) and minute five (8.8 ! 0.75 vs. 8.85! 0.37)

**Table 1.** Demographic characteristics.

|  | Spinal Anesthesia | Epidural Anesthesia |
| --- | --- | --- |
| Number of cases | 195 | 7 |
| Age (years) | $29.8 \pm 5.57$ | $26 \pm 2.16$ |
| Nulliparous | 53 | 1 |
| Multiparous | 142 | 6 |
| ASA |  |  |
| I | 58 (29.7%) | 3(42.8%) |
| II | 134 (68.7%) | 4 (57.2%) |
| III | 3 (1.6%) | 0 (0%) |

Data is represented as frequency (%) or mean ± SD. Source: Data collected from the HGPS database

**Table 2.** Perioperative Events.

|  | Spinal Anesthesia | Epidural Anesthesia |
| --- | --- | --- |
| Total anesthesia time (min) | 77.1 ± 13.06 | 85 ± 10.41 |
| Hipotensión | 123 (63.0%) | 3 (42.8%) |
| Efedrina | 120 (61.5%) | 3 (42.8%) |

Data is represented as frequency (%) or mean ± SD. Source: Data collected from the HGPS database

**Table 3.** Postoperative Events.

|  | Spinal Anesthesia | Epidural Anesthesia |
| --- | --- | --- |
| APGAR (1 min) | 7.81± 0.68 | 7.85 ± 0.37 |
| APGAR (5 min) | 8.8 ± 0.75 | 8.85 ± 0.37 |
| Post dural puncture headache | 6 (3.07%) | 0 (0%) |

Data is represented as frequency (%) or mean ± SD. Source: Data collected from the HGPS database

**Table 4.** Results of the Student’s t test and the Chi square test.

|  | *p* | *OR* | *CI 95%* |
| --- | --- | --- | --- |
| Total anesthesia time (min) | <0.0001 |  |  |
| Hipotension | <0.0001 | 2,25 | 1.51- 3.35 |
| Ephedrine | 0,0004 | 2,11 | 1.41-3.19 |
| APGAR (1 min) | 0,30 |  |  |
| APGAR (5 min) | 0,38 |  |  |
| Post dural puncture headache | 0,06 |  |  |

OR= odds ratio; CI= confidence interval. Source: Table 2 and 3

## DISCUSSION

Regional anesthesia, regardless of the technique used, has been shown to be superior to the use of general anesthesia for scheduled cesarean delivery.^11^ In this study, we focused on patients who received an indication for cesarean section and in whom regional anesthesia was used. A total of 202 patients were studied, of whom only 7 received epidural anesthesia and the rest, that is, the majority, received spinal anesthesia. These data suggest that the most common anesthesia technique used at the General Plaza de la Salud Hospital, HGPS, is spinal anesthesia. International studies suggest similar results, demonstrating that spinal anesthesia is the anesthetic technique of choice used in cesarean deliveries. This is for reasons such as its fast induction time, its empirical association with fewer side effects, and the fact that many anesthesiologists prefer it based on their experiences.^6^

Regarding the demographic data, the mean age of the patients was 29.8 ! 5.57 in those who received spinal anesthesia and 26 ! 2.16 in those who received epidural anesthesia. Of the women who received spinal anesthesia, 27.2% were nulliparous (n=53) and 72.8% were multiparous (n=142). Of the women who received epidural anesthesia, 14.3% were nulliparous (n=1) and 85.7% were multiparous (n=6). Finally, regarding the ASA classification for the spinal group, 29.7% (n=58) were ASA I, 68.7% (n=134) were ASA II, and 1.6% (n=3) were ASA III. For the epidural group, 42.8% (n=3) were ASA I, 57.2% (n=4) were ASA II, and there were no ASA III. No statistically significant differences were found between the two groups for any of these variables.

Mean total anesthetic time for spinal anesthesia was 77.1 ! 13.06. Mean total anesthetic time for epidural anesthesia was 85 ! 10.41 (p=<0.0001). From this we can conclude that spinal anesthesia is associated with shorter total anesthetic time compared to epidural anesthesia. Our findings are similar to those of the study by Ng et al. (2004), in which they concluded that spinal anesthesia generally provides a faster induction time and shorter total anesthetic time compared to epidural anesthesia. The study by Huang et al. (2015) also presented similar results, and concluded that this is one of the reasons why anesthesiologists prefer to use spinal rather than epidural anesthesia in most cases, On the other hand, in our study, the group that received spinal anesthesia presented a greater decrease in blood pressure, so a higher rate of ephedrine administration was seen in these patients. In the spinal anesthesia group, 63.0% of the patients presented episodes of hypotension in which the systolic pressure decreased >20%. While in the epidural anesthesia group, this only occurred in 42.8% of the patients (p=<0.0001). In the same way, a greater use of ephedrine was seen in patients undergoing spinal anesthesia (61.5% vs 42.8%, p=0.0004). Our findings are similar to those of the study by Huang et al. (2015), in which spinal anesthesia had a shorter total anesthetic time than epidural, however more treatment was required for hypotension. All this agrees with the literature that establishes that the spinal technique may be accompanied by unwanted effects such as hemodynamic changes, in this case, hypotension despite resorting to prophylactic measures such as uterine displacement and prehydration. (Lacassie, 2021)

When performing the Chi square test with the collected hypotension data, an odds ratio of 2.25 was seen with a 95% confidence interval of 1.51-3.35. This means that the probability of presenting an episode of hypotension (dysfunction of systolic pressure >20%) is 2.25 times greater in the group of patients in which spinal anesthesia was used. Similarly, for the use of ephedrine, an odds ratio of 2.11 was calculated with a 95% confidence interval of 1.41-3.19. This denotes that patients in the group receiving spinal anesthesia were 2.11 times more likely to be given ephedrine compared to the group receiving epidural anesthesia.

Regarding the secondary outcomes of our study, there was no difference in neonatal outcome between the two groups. In this study, neonatal outcome was evaluated by comparing APGAR scores in newborns at 1 and 5 minutes of life. For the APGAR at minute 1, the means were 7.81 ! 0.68 vs. 7.85 ! 0.37 for spinal and epidural respectively (p= 0.30). For the APGAR at minute 5, the means were 8.8 ! 0.75 vs. 8.85 ! 0.37 for spinal and epidural respectively (p= 0.38). These results are compared to those presented in the study by (Huang et al., 2015) in which they likewise established that there was no association between the regional anesthesia technique used and neonatal outcome.

In addition, 6 cases of post-puncture headache were reported after spinal anesthesia, and none after epidural anesthesia. Parturients who received spinal rather than epidural anesthesia had an additional risk of developing post-puncture headache, although the incidence was extremely low and no statistically significant difference was demonstrated (p= 0.06). This is probably due to the fact that the sample is not large enough to demonstrate the association between both variables; however, the literature indicates that spinal anesthesia has a higher rate of post-puncture headache in patients. (Choi et al., 2018)

Certain limitations were found in this study, these are due to the fact that data such as the anesthesia failure rate, the time from anesthesia induction to surgical incision, the administration of sedatives or analgesics, the dose of morphine administered, scores of the analog scale of pain on the first postoperative day and maternal satisfaction, which due to the scarcity of this information in the clinical records of the HGPS patients could not be collected and analyzed for this study. If it could have been studied, data such as these provide more information to help determine which of the regional anesthesia techniques would be the most appropriate to use based on how one of them is associated with fewer adverse effects compared to the other.

In conclusion:

- The most common anesthetic technique used at the General Plaza de la Salud Hospital, HGPS is spinal anesthesia.
- In terms of demographics, there is no difference in age, parity, or ASA among patients receiving epidural or spinal anesthesia.
- Spinal anesthesia is associated with shorter total anesthetic time compared to epidural anesthesia.
- Spinal anesthesia is associated with greater hemodynamic changes and ephedrine administration compared with epidural anesthesia.
- Neonatal outcomes were similar for both groups.
- Parturients who received spinal rather than epidural anesthesia had an additional risk of developing post-puncture headache, although the incidence was extremely low and no statistically significant difference was shown.
- Both anesthetic techniques have their advantages and disadvantages, so it is difficult to really identify the anesthetic technique associated with better patient outcomes.

### RECOMMENDATIONS

After an exhaustive analysis of the collected data and its comparison with the bibliographic precedents, the following recommendations are urged:

- Carrying out a prospective study in which the anesthetic technique to be used can be assigned equitably and randomly. In this way there will be greater control over the data to be collected based on the already predetermined variables and causality can be established based on the results.
- Expand the sample of the study, turning it into a national and later international study that helps to expand data that suggests which of the regional anesthesia techniques is associated with better results.
- To investigate if there are or are the criteria or guidelines taken into account by anesthesiologists, both specialists and in residency programs, to choose one anesthetic technique over the other.
- Future studies could include more data on neonates who subsequently required treatment as another of the postoperative effects to study.
- Other studies would allow further investigation of other elements that are part of the perioperative effects, such as the effects of the administration of opioid drugs.

## Data Availability

All data produced in the present study are available upon reasonable request to the authors

## Transparency Declaration

I, the lead author (the manuscript’s guarantor) affirm that the manuscript is an honest, accurate, and transparent account of the study being reported; that no important aspects of the study have been omitted; and that any discrepancies from the study as planned (and, if relevant, registered) have been explained.

## Details of Ethical Approval

The research was submitted and approved by the Research Ethics Committee of the Universidad Iberoamericana (UNIBE). It is led by the Dean of Research and Innovation (DEII) and is in charge of evaluating research projects with human beings to ensure that they are carried out responsibly and safely, adhering to international ethical standards.

## Patient and public involvement statement

Patients weren’t approached directly; the data for this study was collected directly from patient files. The personal data of the patients who participated in the study, such as the name, address and telephone number will not be published in order to ensure the protection of the rights of human participants who participate in the research and protect their privacy and integrity.

## Discussion Points

This study compares the two regional anesthetic techniques, spinal and epidural used in cesarean patients. We compared their perioperative results, their postoperative results and compared which technique is associated with better results in both the patients and the product.

## ANNEXES

### DATA COLLECTION INSTRUMENT

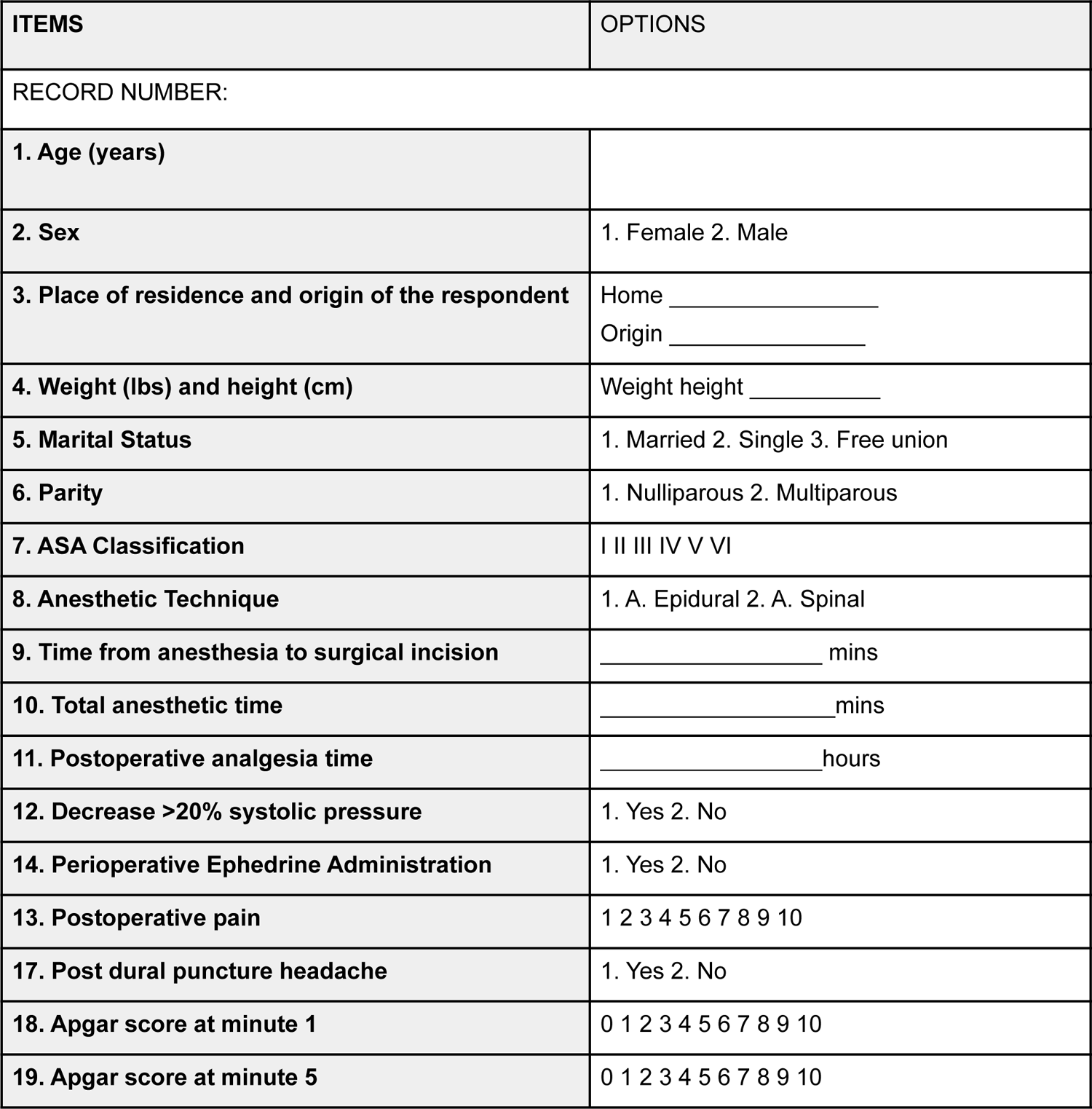

### Schedule of activities

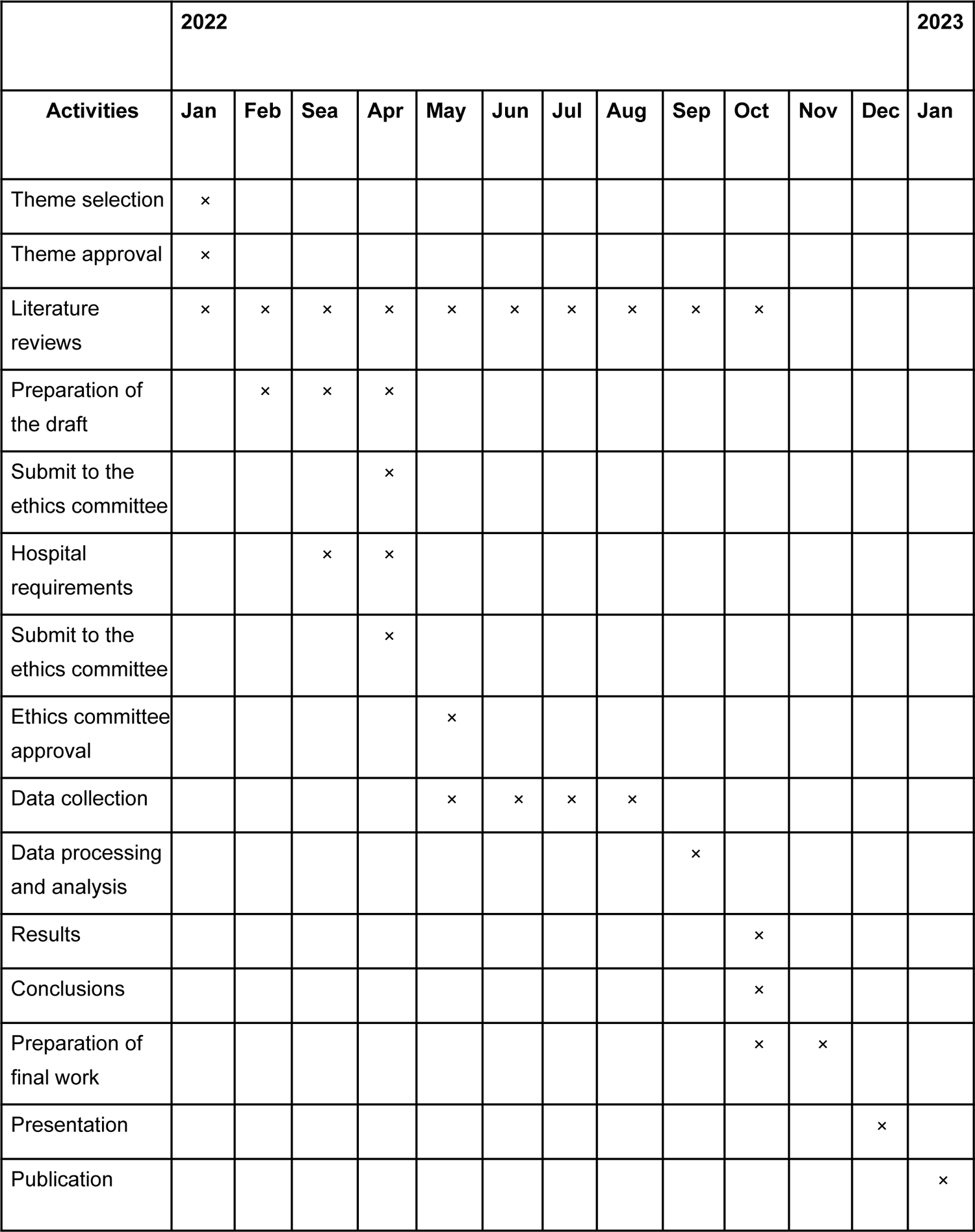

### Budget

| Description | Spent |
| --- | --- |
| Photocopies | US\$ 40.00 |
| Transport | US\$ 100.00 |
| Miscellaneous expenses (non-specific) | US\$ 50.00 |

